# Parapneumonic effusions related to *Streptococcus pneumoniae*: serotype and disease severity trends from 2006 to 2018 in Bristol, UK

**DOI:** 10.1101/2022.03.16.22272461

**Authors:** Catherine Hyams, David T Arnold, Robyn Heath, Zahin Amin-Chowdhury, David Hettle, Gabriella Ruffino, Paul North, Charli Grimes, Norman K Fry, Philip Williams, Robert Challen, Leon Danon, O Martin Williams, Shamez Ladhani, Adam Finn, Nick A Maskell

## Abstract

**Rationale:** *Streptococcus pneumoniae* epidemiology is changing in response to vaccination and some data suggest empyema incidence is increasing. However, differences exist between UK and USA studies. We describe trends in the clinical phenotype of adult pneumococcal pleural infection, including simple parapneumonic effusions (SPE) in the pneumococcal conjugate vaccination (PCV) era.

**Objectives:** To determine whether there were differences in pneumococcal disease presentation and severity associated with pleural infection.

**Methods:** A retrospective cohort study, all adults ≥16 years admitted to three large UK hospitals, 2006-2018 with pneumococcal disease. 2477 invasive pneumococcal cases were identified: 459 SPE and 100 pleural infection cases. Medical records were reviewed for each clinical episode. Serotype data were obtained from the UK Health Security Agency national reference laboratory.

**Results:** Incidence increased over time, including non-PCV-serotype disease. PCV7-serotype disease declined following paediatric PCV7 introduction, but the effect of PCV13 was less apparent as disease caused by the additional six serotypes plateaued with serotypes 1 and 3 causing such parapneumonic effusions from 2011 onwards.

Patients with pleural infection had a median survival 468 days (95% CI 340-590), versus 286 days (95% CI 274-335) in those with SPE. Pleural infection associated with frank pus had lower 90-day mortality than pleural infection without pus (0% versus 29%, *P*<0.0001). 90-day mortality could be predicted by baseline increased RAPID (Renal, Age, Purulence, Infection source, and Dietary factors) score (HR15.01, 95% CI 1.24-40.06, *P*=0.049).

**Conclusions:** Pneumococcal infection continues to cause severe disease despite the introduction of PCVs. The predominance of serotype 1 and 3 in this adult UK cohort is in keeping with previous studies in paediatric and non-UK studies. Rising non-PCV serotype disease and limited impact of PCV13 on cases caused by serotypes 1 and 3 offset the reductions in adult pneumococcal parapneumonic effusion disease burden observed following introduction of the childhood PCV7 programme.

**KEY MESSAGES:** *What is already known on this topic:* The epidemiology of pneumococcal infection is changing in both adults and children following pneumococcal conjugate vaccine (PCV) introduction, as a result of direct and indirect vaccine effects. Other studies have reported that serotypes 1 and 3 disproportionately cause pneumococcal pleural disease; however, the clinical phenotype of parapneumonic effusions associated with pneumococcal infection in adults following PCV introduction is not well described.

*What this study adds:* In this study which presents the largest cohort of patients with a single-organism pleural infection, we demonstrate an increasing incidence of parapneumonic effusions related to *Streptococcus pneumoniae* in adults, attributable to serotype 1 and 3 disease, despite the introduction of PCV13 in the UK childhood vaccination programme. Interestingly, our data suggest that pneumococcal pleural infection is associated with improved survival up to one-year compared to patients with pneumococcal simple parapneumonic effusions.

*How this study might affect research, practice or policy:* Careful assessment of the need for specialist respiratory and thoracic surgical intervention in the context of increasing incidence of adult parapneumonic effusions related to *Streptococcus pneumoniae* will be required, in addition to ongoing monitoring of the effect on serotype distribution and clinical phenotype of current and future vaccines against pneumococcus.

## INTRODUCTION

*Streptococcus pneumoniae* remains the leading bacterial cause of community-acquired pneumonia (CAP), despite vaccine deployment. Approximately 15-20% hospitalised pneumococcal CAP cases are associated with a pleural effusion^1 2^ and 6% with empyema^1 3^. The morbidity and mortality associated with simple parapneumonic effusion (SPE) are higher than with uncomplicated pneumonia^4^. Several USA studies have shown that pneumococcal conjugate vaccine (PCV, a vaccine comprised of pneumococcal polysaccharides conjugated to a non-toxic diphtheria protein) introduction resulted in a significant reduction in both carriage and invasive pneumococcal disease (IPD) due to vaccine serotypes, especially in children^5 6^. Increasing pneumococcal empyema rates in children have been reported^7^ although the effect of paediatric PCV deployment on adult disease phenotype is debated^8^. Recent evidence suggests that incidence of pneumococcal empyema, especially in adults aged >65 years, has increased following paediatric PCV deployment, possibly due to serotype 1 or 3 emergence^9^. Thus, vaccine-driven serotype replacement may be leading to changes in pneumococcal disease phenotype and severity.

A dose of PPV-23 (unconjugated 23-valent polysaccharide vaccine) is offered to all adults ≥65 years in the UK, and ≥2-year-olds with at least one clinical risk factor for pneumococcal disease also receive this vaccine^10^. A 7-valent PCV was introduced into the UK childhood vaccination programme in 2006, subsequently replaced in April 2010 with a 13-valent PCV (PCV13) and the schedule was modified in 2020 so that only two vaccine doses are now given (one at 12 weeks of age, followed by a booster at 12-13 months of age). By 2016/17 in the UK, PCV7 serotype pneumococcal disease in children had virtually disappeared and PCV7 disease significantly reduced in adults. In contrast, PCV13 serotype disease has not disappeared, but plateaued, with a remaining residual incidence of 7·97 per 100,000 across all age groups and disease attributable to non-PCV13 serotypes has increased, especially in the elderly^11-13^. Evidence from a large pneumococcal pneumonia patient cohort in Nottingham, UK suggests that, aside from age, residential care status and some comorbidities, there are relatively few differences between patients with respiratory infection caused by PCV13 and those with non-PCV13 serotype infections^14^.

Here we present the largest cohort of patients with a single-organism pleural infection, encompassing simple parapneumonic effusion (SPE)^15^ and pleural infection (complex parapneumonic effusion (CPE) or empyema)^3 15^ attributable to pneumococcus, in this single-centre observational study covering thirteen years during which PCVs were introduced into the UK paediatric vaccination programme. We sought to determine the incidence of parapneumonic effusions in adults, overall and by vaccine-serotype group. Additionally, we aimed to describe differences in pneumococcal disease presentation and severity associated with pleural infection, given concerns surrounding changing pneumococcal serotype distribution and serotype 1 and 3 pleural infection.

## MATERIALS AND METHODS

### Study Subjects

Patients aged ≥16 years admitted to all three hospitals providing emergency care in Bristol and Bath (University Hospitals Bristol and Weston, North Bristol and The Royal United Hospital NHS Trusts) between January 2006 and December 2018, with a confirmed microbiological diagnosis of pneumococcal infection were eligible for this study. This study was approved by the Health Research Authority, UK (IRAS 265437).

### Study Design

A retrospective cohort study at three large NHS hospitals in the UK.

### Patient and public involvement

There was no patient or public involvement in the undertaking of this study.

### Methods

Study-eligible cases were identified retrospectively by searching the Laboratory Information Management System (LIMS) database (Clinisys WinPath Enterprise). *S. pneumoniae* was confirmed on culture from sterile-site sampling at a central laboratory using standard microbiological techniques combined with API®-20 Strep (BioMérieux, UK) or MALDI-TOF (matrix-assisted laser desorption/ionisation/time of flight) mass spectrometry (Bruker, UK) ^16^. A positive pneumococcal urinary-antigen test (BinaxNOW®, Alere, UK) was also considered confirmative of pneumococcal infection. Patients were included if they tested positive on either or both tests. Any patient with pleural fluid which cultured a single-organism which was not pneumococcus was excluded from this cohort; although, multi-organism pleural infection including pneumococcus was included.

Confirmed cases were linked with the UK Health Security Agency (UKHSA) national reference laboratory to obtain serotype data ^17^ which were collected separately from clinical data to avoid any risk of bias in data collection. Pneumococcal serotypes were grouped by PCV (PCV7; PCV13-7 representing the additional 6 serotypes in PCV13; and PCV13) or as non-PCV (Supplementary Data 1).

Patients with pneumococcal infection and a pleural effusion on radiology were either classified as:

1. pleural infection: if pleural fluid was pus or bacterial culture positive (i.e. an empyema)^3^; an effusion relating to a current pneumonia episode which necessitated definitive drainage (e.g. thoracic surgery); or, there was an exudative effusion^18^ with pH ≤7.2^3^; or,
2. SPE: an exudative effusion not classified as pleural infection in a patient with a clinical and/or radiological diagnosis of pneumonia^15^.

Patients’ clinical records were reviewed at each hospital and data, including clinical outcomes, recorded. The vaccination status of each patient was established from electronically linked GP records^16^. The CURB65 severity score on admission was calculated for each clinical episode^19^. Patients with pleural infection had a RAPID (Renal, Age, Purulence, Infection source, and Dietary factors) score calculated^20^, which has been validated to stratify adults with pleural infection according to increasing risk of mortality^21^.

### Study objectives

The primary objective was to determine trends in pneumococcal pleural disease incidence between 2006 and 2020, both overall and by vaccine-serotype groups, in the context of the UK vaccination programme. Secondary objectives included determining if any changes in disease incidence were attributable to emergence of either vaccine-serotype groups or specific serotypes, as well as describing differences in pneumococcal disease presentation and disease severity associated with pleural infection.

### Analysis

Patient data are reported as medians and interquartile ranges (IQR) for continuous variables, or means and standard deviations where their distribution was confirmed to be normal using the Anderson Darling normality test. Categorical variables are presented as counts and percentages. Baseline characteristics and comorbid risk factors for SPE and pleural infection were compared using Fisher’s exact tests for categorical variables, and the two-sample Wilcoxon Rank Sum test for non-parametric continuous variables, or 2 sided student’s t-test for parametric continuous variables. Comparisons were not performed where missing data was present. *P*-values are presented unadjusted but the level at which a result was considered significant was reduced in recognition of the fact multiple comparisons were made. Patient survival was assessed at two years (730 days) following hospital admission, with this being the time of right censoring, and a median survival calculated. Time series trends for serotype proportions were estimated using a 5-knot natural spline, fitted to a quasi-binomial model, using a logistic link function in a maximum likelihood estimation framework, The Kaplan– Meier (KM) method and Cox proportional hazard regression model were used for calculation of survival at 30-, 90- and 365-days following first positive pneumococcal microbiological test. Proportional hazard assumptions were tested by visual assessment of the KM survival curve, log(-log) plots and Schoenfeld residuals. Survival differences were analyzed using the log-rank test. Statistical analysis was performed using SPSS, version 28.0 (New York, IBM) or with R v4.0.2. Graphs were generated in GraphPad PRISM, version 9.0.

## RESULTS

2657 episodes of pneumococcal infection were identified, of which 2447 were respiratory infections: 1888 pneumonia-only cases, 459 with SPE and 100 cases including pleural infection (Supplementary Data 2). In total, 282/559 (50%) were male with a median age of 68y (IQR 50-78) (Table 1). Patients with pleural infection were younger than those with SPE (median ages 55 vs 71y respectively, *P*<0.001), were more likely to have history of cardiac disease (*P*<0.015) and less likely to develop bilateral effusions (*P*<0.001). On presentation to hospital, neither clinical observations nor CURB65 score differed significantly between patients with SPE and pleural infection (*P*=0.024); in contrast, patients with pleural infection had significantly greater C-reactive protein levels, and other inflammatory markers including white cell count and neutrophil count were tending towards elevation (Table 1).

**Table 1:** Clinical features of patients with pneumococcal parapneumonic effusions.

| Variable | Characteristic | All parapneumonic effusions | SPE | Pleural infection | P value * |
| --- | --- | --- | --- | --- | --- |
|  |  | Value (N=559) | Value (N=459) | Value (N=100) |  |
| Streptococcus pneumoniae diagnosis |  |  |  |  |  |
| Micro confirmation** | Blood culture % (N) | 50.6% (283) | 52.1% (239) | 44.0% (44) | 0.15 |
|  | Urine antigen test % (N) | 49.4% (276) | 47.9% (220) | 56.0% (56) |  |
| Pleural culture | Negative % (N) | 92.5% (517) | 100.0% (459) | 58.0% (58) | <0.001 |
|  | S. pneumoniae % (N) | 4.8% (27) | 0.0% (0) | 27.0% (27) |  |
|  | Multi-organism % (N) | 2.7% (15) | 0.0% (0) | 15.0% (15) |  |
| Demographics and Comorbidities |  |  |  |  |  |
| Gender | Male % (N) | 50.4% (282) | 50.1% (230) | 52.0% (52) | 0.74 |
|  | Female % (N) | 49.6% (277) | 49.9% (229) | 48.0% (48) |  |
| Age (years) | Median [IQR] | 68 [50—80.5] | 71 [54—82] | 55 [45—73.2] | <0.001 |
| Smoking status | Non-smoker % (N) | 22.9% (128) | 23.1% (106) | 22.0% (22) | 0.0068 |
|  | Current smoker % (N) | 35.1% (196) | 32.2% (148) | 48.0% (48) |  |
|  | Ex-smoker % (N) | 42.0% (235) | 44.7% (205) | 30.0% (30) |  |
| Chronic lung disease | Rate % (N) | 43.8% (245) | 44.2% (203) | 42.0% (42) | 0.74 |
| Cardiac disease | Rate % (N) | 53.3% (298) | 55.8% (256) | 42.0% (42) | 0.015 |
| Upper GI | Rate % (N) | 38.3% (214) | 39.2% (180) | 34.0% (34) | 0.36 |
| Cerebrovascular disease | Rate % (N) | 7.0% (39) | 7.4% (34) | 5.0% (5) | 0.52 |
| Type 2 Diabetes | Rate % (N) | 17.0% (95) | 17.4% (80) | 15.0% (15) | 0.66 |
| Chronic Liver Failure | Rate % (N) | 4.5% (25) | 4.6% (21) | 4.0% (4) | 1 |
| Intravenous Drug Usage | Rate % (N) | 4.3% (24) | 3.7% (17) | 7.0% (7) | 0.17 |
| Alcohol excess | Rate % (N) | 9.5% (53) | 8.3% (38) | 15.0% (15) | 0.057 |
| BMI ≤17 or ≥35 | Rate % (N) | 4.1% (23) | 3.3% (15) | 8.0% (8) | 0.047 |
| CURB-65 Score |  |  |  |  |  |
| CURB-65 on admission | 0 % (N) | 24.5% (137) | 22.4% (103) | 34.0% (34) | 0.024 |
|  | 1 % (N) | 24.5% (137) | 24.4% (112) | 25.0% (25) |  |
|  | 2 % (N) | 26.1% (146) | 28.3% (130) | 16.0% (16) |  |
|  | 3–5 % (N) | 24.9% (139) | 24.8% (114) | 25.0% (25) |  |
| Laboratory Tests and Radiological Features |  |  |  |  |  |
| White cell count, ×10<sup>9</sup>/L | Mean ± SD | 17.9 ± 8.19 | 17.5 ± 8.02 | 19.9 ± 8.71 | 0.011 |
| Neutrophil count, ×10<sup>9</sup>/L | Median [IQR] | 14.9 [10—20.2] | 14.2 [9.9—20.1] | 17.9 [10.4—23.9] | 0.0028 |
| C-reactive protein, mg/dL | Median [IQR] | 215 [132—341] | 200 [129—312] | 338 [204—412] | — |
| Albumin, g/dL | Median [IQR] | 30 [26—36] | 31 [27—36] | 28 [23—34] | <0.001 |
| Urea, mmol/L | Median [IQR] | 9.3 [6.4—13.2] | 9.2 [6.5—13.3] | 9.8 [5.97—12.8] | 0.67 |
| Bilateral effusion | Rate % (N) | 24.9% (139) | 28.1% (129) | 10.0% (10) | <0.001 |
| Outcomes |  |  |  |  |  |
| Acute Renal Failure† | Rate % (N) | 35.4% (198) | 37.3% (171) | 27.0% (27) | 0.064 |
| Liver dysfunction# | Rate % (N) | 10.7% (60) | 12.0% (55) | 5.0% (5) | 0.048 |
| ITU admission | Rate % (N) | 20.2% (113) | 20.3% (93) | 20.0% (20) | 1 |
| Increased care needed | Rate % (N) | 35.4% (198) | 38.3% (176) | 22.0% (22) | 0.0018 |
| Admission days | Median [IQR] | 11 [6—20] | 10 [5—19.5] | 15 [10—21] | <0.001 |
| **Mortality** |  |  |  |  |  |
| Inpatient mortality | Rate % (N) | 12.7% (71) | 13.1% (60) | 11.0% (11) | 0.74 |
| 30-day mortality | Rate % (N) | 13.2% (74) | 14.2% (65) | 9.0% (9) | 0.19 |
| 90-day mortality | Rate % (N) | 19.0% (106) | 20.5% (94) | 12.0% (12) | 0.05 |
| 1-year mortality | Rate % (N) | 27.0% (151) | 29.2% (134) | 17.0% (17) | 0.013 |

\* Significance determined using Fisher's exact test (categorical variables) or 2 sample Wilcoxon Rank Sum test (continuous variables). Normality of distributions determined using Anderson-Darling normality test. Due to the fact multiple comparisons are made we consider the level of significance required to exclude the null hypothesis to be 0.0017 (0.05/30).
\*\* Patients were included if they tested positive for pneumococcus on sterile site culture and/or urinary antigen test. Therefore some patients tested positive for both tests.
† Acute renal failure was defined as a reduction in renal function (as measured by creatinine or eGFR), oliguria (<400ml urine per 24h) or clinical diagnosis of acute kidney injury

Annual incidence of *S. pneumoniae* parapneumonic effusions increased throughout the study period (Figure 1A) with a 33% and 34% increase in unplanned hospital admissions and microbiological testing between 2006 and 2018 (Supplementary Data 4), indicating relative increase in hospital workload was not equivalent to the increase in disease incidence. The age of patients admitted throughout the study did not show any apparent trend (Supplementary Data 5). In 38% cases serotype was available, of which overall 52% were attributable to a PCV13 serotype while only 8% were caused by a non-vaccine serotype. There was an absolute and proportional decrease in disease caused by PCV7 serotypes (Figure 1). PCV13-7 disease showed an initial rise, followed by a fall after PCV13 introduction with a gradual rise in disease over the last 5 study years (Figure 1B). The only PCV13-7 serotypes that caused pneumococcal pleural infections after 2010 were 1 and 3: 2 (17% of serotype known disease) and 3 (25%) pre-2010 pleural infection cases versus 9 (30%) and 7 (23%) post-2010 pleural infection cases due to serotype 1 and 3 respectively (Supplementary Data 6). Serotype 1 affected younger patients than serotype 3 but was associated with higher RAPID severity group (high RAPID severity group 36% versus 17%, *P*<0.0001) and a trend towards increased rates of renal failure (77% versus 43%, *P*=0.023) than seen in patients with serotype 3 parapneumonic effusions (Table 2). After 2011, all PCV7 disease was attributable to serotypes 14 and 19F, with all PCV7 disease being SPE and no cases of pleural infection.

**Figure 1:**
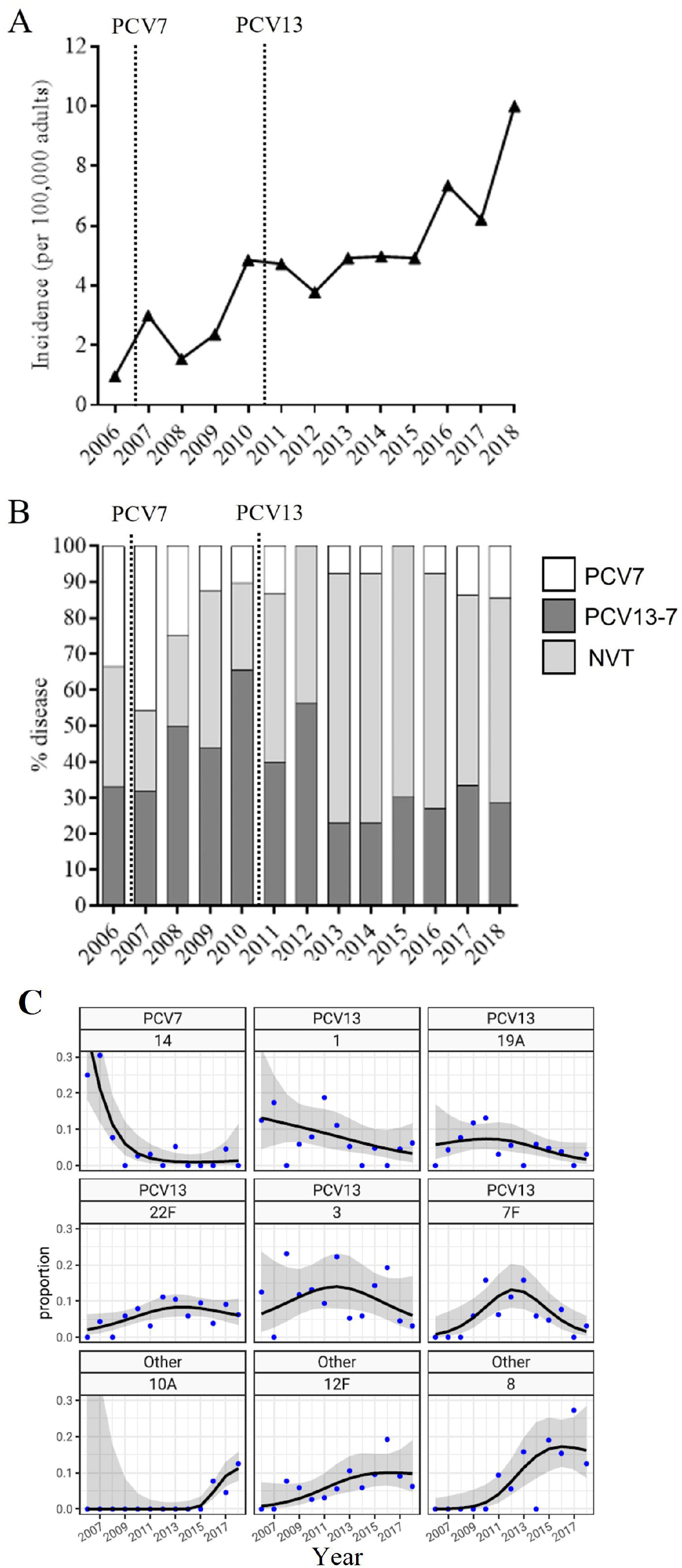
Vaccine group serotype trends in pneumococcal parapneumonic effusions in Bristol, UK 2006-2018. (A) Incidence of pneumococcal parapneumonic effusion (per 100,000 adults), calculated using adult population data from the Office of National Statistics (Supplementary Data 5). (B) proportion of serotyped disease caused by vaccine serotype group as shown from 2006-18. Dashed lines show the introduction of paediatric PCV7 and PCV13. (C) proportion of serotyped disease caused by the major individual serotypes from 2006-18. The black line shows the trend in proportion of disease, with the grey area representing the 95% confidence interval.

**Table 2:** Clinical features of patients with serotype 1 and 3 pneumococcal parapneumonic effusions.

| Variable | Characteristic | Serotype 1 | Serotype 3 | P value * |
| --- | --- | --- | --- | --- |
|  |  | N=22 | N=30 |  |
| Demographics and Comorbidities |  |  |  |  |
| Gender | Male % (N) | 59.1% (13) | 53.3% (16) | 0.78 |
|  | Female % (N) | 40.9% (9) | 46.7% (14) |  |
| Age (years) | Mean ± SD | 53.4 ± 18.9 | 72.5 ± 15.9 | <0.001 |
| Smoking status | Non-smoker % (N) | 27.3% (6) | 23.3% (7) | 0.45 |
|  | Current smoker % (N) | 50.0% (11) | 36.7% (11) |  |
|  | Ex-smoker % (N) | 22.7% (5) | 40.0% (12) |  |
| Chronic lung disease | Rate % (N) | 45.5% (10) | 40.0% (12) | 0.78 |
| Cardiac disease | Rate % (N) | 40.9% (9) | 70.0% (21) | 0.049 |
| Upper GI | Rate % (N) | 13.6% (3) | 36.7% (11) | 0.11 |
| Cerebrovascular disease | Rate % (N) | 0.0% (0) | 10.0% (3) | 0.25 |
| Type 2 Diabetes | Rate % (N) | 18.2% (4) | 23.3% (7) | 0.74 |
| Chronic Liver Failure | Rate % (N) | 0.0% (0) | 6.7% (2) | 0.5 |
| Intravenous Drug Usage | Rate % (N) | 4.5% (1) | 3.3% (1) | 1 |
| Alcohol excess | Rate % (N) | 9.1% (2) | 13.3% (4) | 1 |
| Pleural Features |  |  |  |  |
| Pleural infection | Rate % (N) | 50.0% (11) | 33.3% (10) | 0.26 |
| Loculation | Rate % (N) | 22.7% (5) | 10.0% (3) | 0.26 |
| Pus | Rate % (N) | 18.2% (4) | 20.0% (6) | 1 |
| RAPID Group | Low % (N) | 50.0% (11) | 16.7% (5) | <0.001 |
|  | Medium % (N) | 13.6% (3) | 66.7% (20) |  |
|  | High % (N) | 36.4% (8) | 16.7% (5) |  |
| Outcomes |  |  |  |  |
| Acute Renal Failure† | Rate % (N) | 77.3% (17) | 43.3% (13) | 0.023 |
| Liver dysfunction# | Rate % (N) | 9.1% (2) | 3.3% (1) | 0.57 |
| ITU admission | Rate % (N) | 18.2% (4) | 20.0% (6) | 1 |
| Increased care needed | Rate % (N) | 27.3% (6) | 33.3% (10) | 0.76 |
| Admission days | Median [IQR] | 11 [5—20.5] | 10 [4.25—18] | 0.37 |
| Inpatient mortality | Rate % (N) | 4.5% (1) | 23.3% (7) | 0.12 |
| 30-day mortality | Rate % (N) | 9.1% (2) | 20.0% (6) | 0.44 |
| 90-day mortality | Rate % (N) | 13.6% (3) | 36.7% (11) | 0.11 |
| 1-year mortality | Rate % (N) | 18.2% (4) | 46.7% (14) | 0.042 |

\* Significance determined using Fisher's exact test (categorical variables) or 2 sample Wilcoxon Rank Sum test, or student's t-tests (continuous variables). Normality of distributions determined using Anderson-Darling normality test. *P*-values were considered significant if they were under 0.002 (0,05/24).
† Acute renal failure was defined as a reduction in renal function (measured by creatinine or eGFR), oliguria (<400ml urine per 24h) or clinical diagnosis of acute kidney injury

Overall, 19% patients with pneumococcal parapneumonic effusions died within 90-days of presentation (Table 1, Supplementary Table 7). Patients with pleural infections had higher survival rates than those with SPE at 30-, 90- and 365-days (*P*<0.01) (Figure 2A). The mortality rate among patients with pleural infection increased alongside RAPID score severity at 30-, 90- and 365-days (Figure 2B, Table 3A), and a high RAPID score was associated with a median survival of 56 days (20-154 days, 95% CI). Proportional hazards regression analysis supported the earlier finding that more severe RAPID group was associated with increased 90-day (*P*=0.049, 95% CI 1.24-40.06) and 365-day mortality (*P*=0.034, 95% CI 1.09-51.2) (Table 3B).

**Figure 2:**
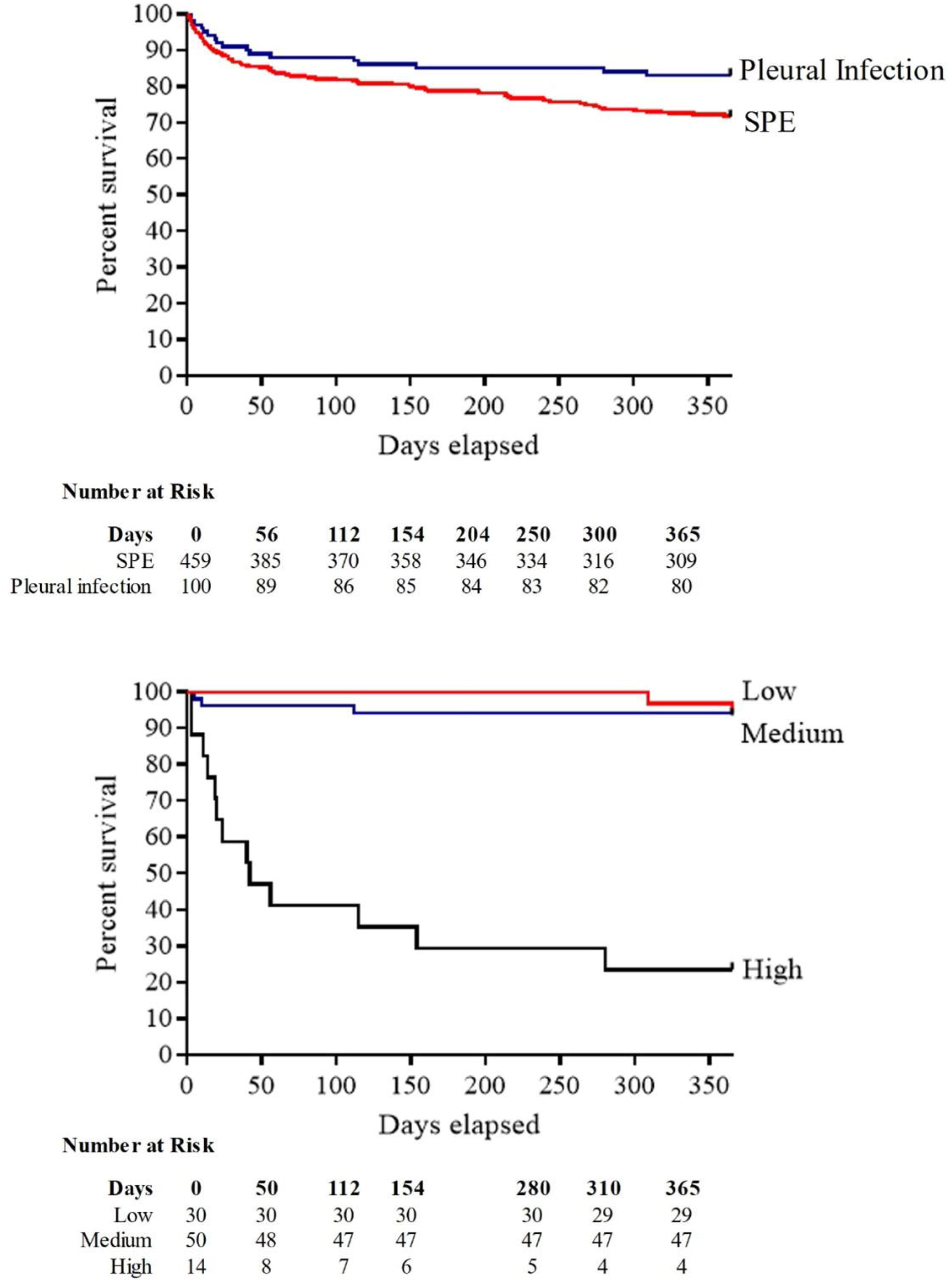
1-year survival in patients with pneumococcal parapneumonic effusions. (A) Kaplan-Meier survival curve and number at risk for patients with simple parapneumonic effusion (SPE) (red line) and pleural infection (blue line). (B) Kaplan-Meier survival curve and number at risk for patients with pneumococcal pleural infection for low (red line), medium (blue line) ad high (black line) RAPID score groups. The number of subjects at risk immediately before each time point is listed in a numbers-at-risk table.

**Table 3:**
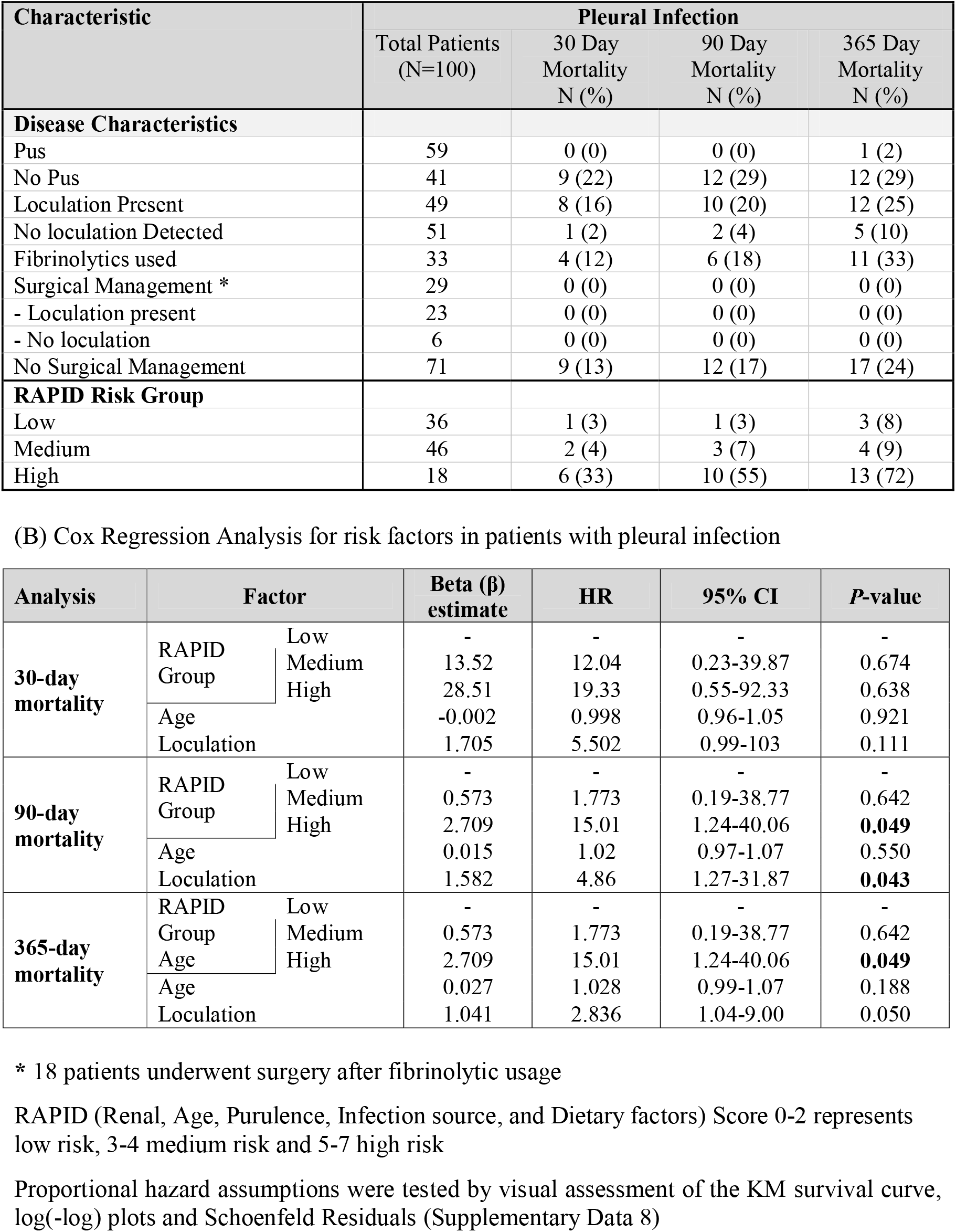
Mortality and Risk Analysis in patients with pleural infection cases. (A) Mortality among 100 pneumococcal pleural infection cases with different clinical features and RAPID severity Scores

Median survival was 468 days (340-590 days, 95% CI) in patients with pleural infection, vs 286 (274-335 days, 95% CI) in those with SPE. In total, 59/100 (59%) pleural infection patients had pus and these patients had improved 30-, 90-, and 365-day mortality compared to those without pus; in contrast, loculation was associated with reduced 30-, 90- and 365-day mortality (Table 3B). Surgical management was associated with improved prognosis: no deaths occurred within 1-year of presentation compared to 24% mortality in patients with pleural infection who did not undergo surgery. Patients undergoing thoracic surgery were younger than those who did not (median age 45 years, IQR40-52 versus 66 years, IQR53-78).

## DISCUSSION

This observational cohort study, with data from a 13-year period spanning PCV introduction into the UK childhood vaccination programme, presents data clearly showing increasing adult pneumococcal parapneumonic effusion disease incidence. The proportion of disease attributable to PCV7 serotypes fell after this vaccine was introduced into the UK childhood vaccination programme; however, disease attributable serotypes 1 and 3 did not show decline following PCV13 vaccine rollout. Further increase in non-vaccine serotype disease contributed to increasing disease incidence. Both overall admissions and microbiological testing at the study hospitals increased progressively during our study period, but not as much as disease incidence, suggesting these did not solely account for the increasing incidence observed. An increasing incidence of pneumococcal disease has also been reported by other UK studies^14 17 22 23^, with Pick *et al* reporting incidence of pneumococcal pneumonia of 32.2 and 48.2 per 100,000 population in 2013 and 2018 respectively ^14^. Using previously published estimates that up to 20% of adults hospitalised with pneumococcal pneumonia develop a pleural effusion^1 2^, estimates of parapneumonic effusion incidence from that study would be 6.4 and 9.6 in 2013 and 2018, which are similar to the incidence rates of 5.0 and 9.8 observed in the same years in this study. Recent national epidemiological studies of pleural infection from North America also show significant increases in pleural infection rates both in children^24^ and adults^25^. Likewise, a Danish study from 1997-2001, showed steadily rising pleural infection rates in the elderly^26^ and a UK study showed annual rises in adult pleural infections from 2008-17^27^, most marked in over 60-year-old individuals with a 194% increase over the decade.

The introduction of PCV7 into the UK childhood vaccination programme in 2006 was followed by a decrease in adult pleural effusions attributable to PCV7 vaccine serotypes in this study. Notably, all parapneumonic effusions due to PCV7 serotypes after 2011 were SPE and due to serotypes 14 and 19F, with no cases of pleural infection. After 2010, all PCV13 pleural infection cases were attributable to serotypes 1 or 3, highlighting the predilection of these serotypes for causing pleural disease. The inclusion of patients with a positive pneumococcal urinary-antigen test permitted serotype identification although in only 38% of cases, despite linkage with the UK Health Security Agency (UKHSA) national reference laboratory. Nevertheless, the trends in serotype and vaccine-serotype group that we found in this cohort are similar to those reported by other surveillance studies. Additionally, the trends in serotype distribution partially align with national serotype trends, showing reductions in both PCV7 serotype and serotype 1 adult IPD incidence^11^, and both serotypes are prevalent in studies of pleural infection in children^28^. In the UK, large reductions in vaccine-serotype adult IPD were offset by increases in non-PCV13 IPD^11 17 22^, a trend also apparent in this patient cohort, with non-PCV13 disease increasing from 30% to 60% of known serotype disease. Pick *et al* showed an increase in non-PCV13 serotype pneumococcal pneumonia from 2014/15 onwards^14^ as well as an overall increase in PCV13-7 serotype disease, in contrast to trends reported in this study. However, Pick *et al* report all respiratory infection and therefore as these are the first data on serotype trends in adult pneumococcal pleural disease since paediatric PCV introduction in the UK, differences reported here may be attributable to differences within patients with pleural infection. This comparison with other UK surveillance studies provides some reassurance that the results here are representative of parapneumonic effusions related to *Streptococcus pneumoniae*. This highlights the difficulty in undertaking accurate disease surveillance, which cannot be conducted without thorough microbiological testing of patients. However, such surveillance is critical for monitoring both the direct and indirect effects of current pneumococcal vaccinations and predicting the effect of novel vaccines before implementation.

Serotype affects pneumococcal resistance to innate immunity, duration of nasopharyngeal colonisation, the number of episodes of invasive disease per colonisation event, mortality and disease severity^29-31^. The two PCV13-7 serotypes which continued to cause pneumococcal pleural infection following PCV13 introduction are notable serotypes as they have unusual polysaccharide capsules. Serotype 1 expresses a zwitterionic and structurally diverse polysaccharide capsule, which uniquely may act as a T-cell dependent antigen^32^, is rarely isolated in asymptomatic nasopharyngeal colonization but is commonly isolated in IPD^29^, frequently causing invasive pneumonia^33^. Serotype 1 is more likely to be identified in young patients, and some studies find it is the most frequent cause of pneumococcal pleural infection and responsible for increasing disease incidence^2 9^. Interestingly, in this cohort, patients with serotype 1 pleural infection had a better survival rate despite higher RAPID score than those with serotype 3. This may reflect the younger age and lower cardiac disease rate in patients with serotype 1 pleural infection, as well as increased likelihood of surgical intervention in these cases. In contrast, the serotype 3 polysaccharide capsule is large, resulting in a highly mucoid appearance, and is one of a few capsules produced by synthase-mediated synthesis^34^. Serotype 3 also causes considerable disease burden^2 28^, is the most common overall cause of pneumonia in the USA^35^, is highly invasive^29^ and associated with increased mortality^30^. Distinct lineages exist within serotype 3 clonal complex 180, and recent clade distributions shift has led to the emergence and expansion of clade II^36 37^, which now represents 50% serotype 3 IPD in the UK^38^. We could not determine serotype 3 clade grouping in this analysis, but clade II emergence may, at least in part, explain serotype 3 disease persistence.

Interestingly, our data suggest that pneumococcal pleural infection is associated with improved survival up to one-year compared to patients with pneumococcal SPE. We found a high burden of pre-existing medical comorbidity in patients with both SPE and pleural infection, in keeping with a large systematic review which found that 72% of patients with pleural infection had at least one significant comorbidity with high levels of pre-existing cardiac and respiratory diseases^39^. However, patients with SPE had a trend towards a high burden of underlying cardiovascular disease, chronic obstructive pulmonary disease, solid-organ and haematological malignancy (Supplementary Data 3), which may account for differences in survival.

It should be noted that <30% of effusions are known to have more than one aetiology^40^, with many having a cardiac or renal component. Thus, it can be difficult to determine the full aetiology of a pleural effusion, and pneumococcal SPEs may often be due to dual aetiology e.g. infection, causing atrial fibrillation and cardiac failure. In this cohort, pre-existing cardiac disease and bilateral effusions were more common in patients with SPE. Previous data suggest individuals with pneumonia and cardiac failure have higher mortality rates^41^, and bilateral effusions are associated with worse outcomes^41 42^. Hence, whilst some effusions reflect more severe lung inflammation, others may reflect concomitant comorbidity in the host; either factor can account for worse outcome and may explain this finding in our cohort. Further, the high proportion of patients undergoing surgical intervention, which is associated with improved outcomes^43 44^, may have improved patient survival in the pleural infection patients. Of note, in our pleural infection group, no deaths occurred within 1-year of presentation in those who underwent surgery compared to 24% mortality in patients who did not. Although patients who did not undergo surgery were older than those who did, these data highlight the importance of appropriate surgical intervention in patients with pleural infection.

As shown in the initial validation and subsequent cohort studies of the RAPID score^20 21^, a high score was strongly associated with the risk of mortality by 3-months. Study patients in the low-risk group had a 3% (95% C.I 0.7 to 15%) risk of 90-day mortality compared to 57% (95% C.I. 28% to 82% *P*<0.01), for those with a high risk score, while there was much less difference in survival between those in low and medium RAPID groups, as reported by others^21 45^. There was a positive association between pleural fluid purulence and survival: this relationship has been demonstrated in cohorts that were not restricted by causative organism, suggesting it is disease and not pathogen specific^20^.

This study identified patients with pneumococcal infection and parapneumonic effusions within a defined geographical area, covering a population of approximately one million adults, and encompassed 3 large hospitals with 100,000 unplanned adult admissions annually. One of these hospitals is the regional thoracic surgery centre, whilst another has a specialist pleural disease service, possibly increasing the accuracy of the clinical data presented here.

We captured disease and serotype trends over 13 years, spanning PCV introduction. By linking with the UKHSA national reference laboratory we were able to report serotype where it was available. Importantly, the epidemiological data were supported with detailed clinical information for individual patients, including short- and long-term outcomes. However, this study also has limitations in addition to those discussed above. This is a retrospective observational study; therefore, only information documented in clinical records could be included and patients were managed at the discretion of individual physicians. The pneumococcal serotype data was only obtainable through standard-of-care testing and by sterile site culture, as BinaxNOW does not derive serotype. Despite linkage with the national reference service, we were only able to determine the serotype in 38% of pneumococcal cases in this cohort. As a regional study, the findings here may not be representative of other populations; although, our serotype trends are comparable to other UK national and regional reports. We note an expanding total and older adult population in Bath and Bristol during the study, which may affect disease incidence. Additionally, changes in patient or physician treatment preferences, including threshold for referral to secondary care, diagnostic testing and treatment which may have occurred and impacted disease incidence estimates^46^. We included patients who tested positive for pneumococcus using the BinaxNOW® urinary antigen test, which has a known sensitivity of 65% and prolonged positive test result after exposure to pneumococcus^47^. Lastly, different methodologies were used to identify pneumococcal serotypes during the study period; whole genomic sequencing was only routinely used by UKHSA from October 2017, and therefore earlier isolates did not have this performed.

Overall, we found an increasing incidence of pneumococcal parapneumonic effusion in this UK population. The proportion of disease attributable to PCV7 serotypes fell after this vaccine was introduced into the UK childhood vaccination programme; however, disease attributable serotypes 1 and 3 did not show decline following PCV13 vaccine rollout. Further increase in non-vaccine serotype disease contributed to increasing disease incidence. Patients with SPE had reduced one-year survival compared to those with pleural infection, which may be attributable to the burden of underlying pre-existing diseases. We also found reassuring evidence that surgical management was associated with improved patient outcomes in patients with pneumococcal pleural infection. Future research should aim to determine if *S. pneumoniae* serotype is predictive of mortality risk in adults with pneumococcal parapneumonic effusions.

## Supporting information

STROBE

## Data Availability

The data used in this study are sensitive and cannot be made publicly available without breaching patient confidentiality rules. Therefore, individual participant data and a data dictionary is not available to other researchers.

## ACKNOWLEDGEMENTS

The authors would like to acknowledge the research teams at The Royal United, North Bristol and University Hospitals of Bristol and Weston NHS Trusts.

