## Supplementary Data for "Parapneumonic effusions related to *Streptococcus pneumoniae*: serotype and disease severity trends from 2006 to 2018 in Bristol, UK"

**Supplementary Data 1: Serotypes contained within each pneumococcal vaccination and vaccination group.**

| **Vaccination** | **Serotypes** |
|---|---|
| Pneumococcal polysaccharide vaccine, 23 valent (PPV-23, PneumoVax®) | 1, 2, 3, 4, 5, 6B, 7F, 8, 9N, 9V, 10A, 11A, 12F, 14, 15B, 17F, 18C, 19F, 19A, 20, 22F, 23F, 33F. |
| Pneumococcal conjugate vaccine, 7-valent (PCV-7, Prevenar®) | 4, 6B, 9V, 14, 18C, 19F and 23F |
| Pneumococcal conjugate vaccine, 13-valent (PCV-13, Prevenar13®) | 1, 3, 4, 5, 6A, 6B, 7F, 9V, 14, 18C, 19A, 19F, and 23F |
| PCV13-7 serotypes | 1, 3, 5, 6A, 7F, 19A |

**Supplementary Data 2: Study flow diagram**

Results from 3677 microbiology database occurrences of a positive blood or urine antigen test were identified. There were 2657 cases of pneumococcal infection in adults, including 559 patients with parapneumonic effusions.

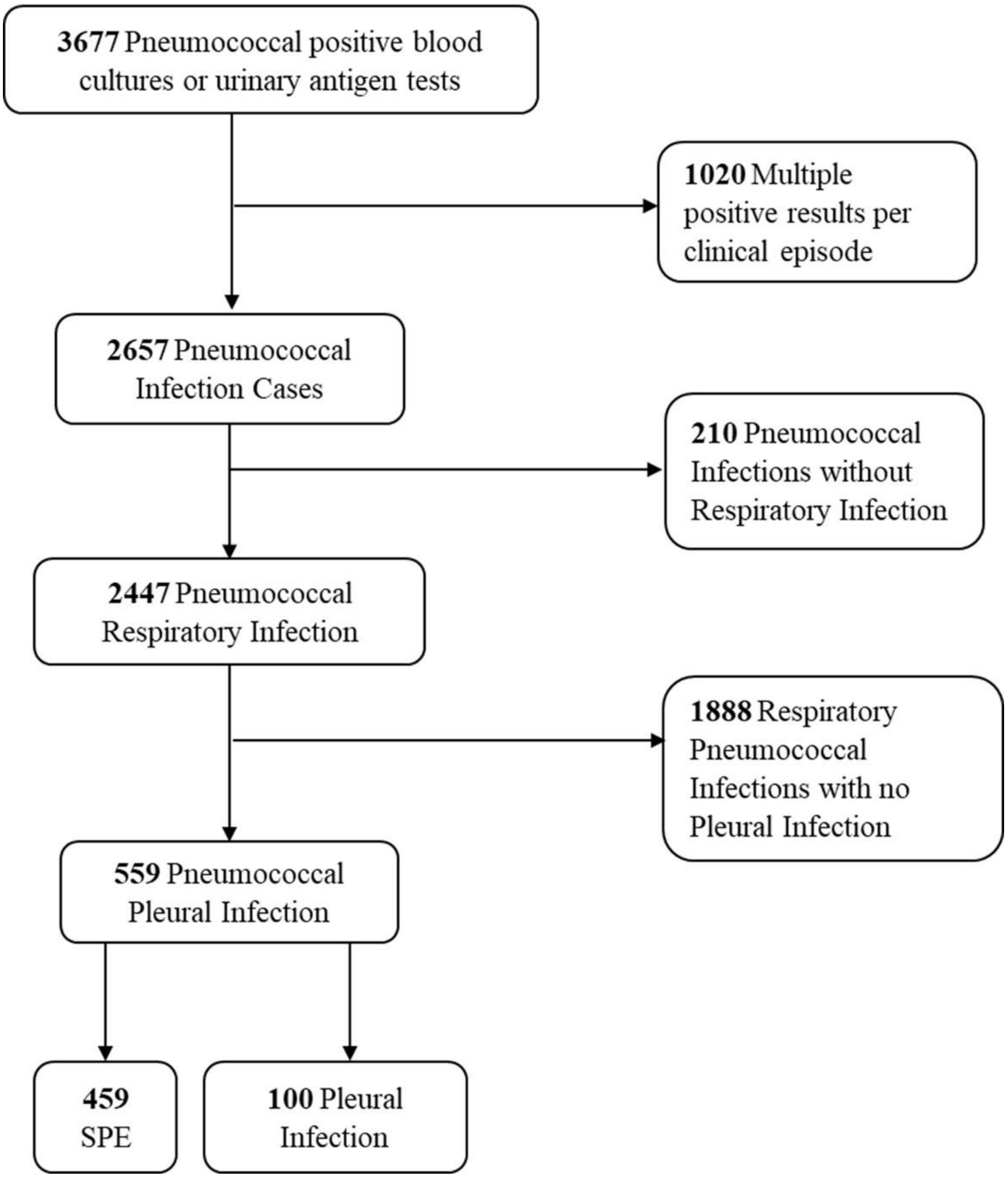

**Supplementary Data 3: Additional characteristics and outcomes of patients admitted with pneumococcal parapneumonic effusions**

| Variable | Characteristic | SPE | Pleural infection | *P* value * |
|---|---|---|---|---|
| | | **N=459** | **N=100** | |
| Index of Multiple Deprivation‡ | Median [IQR] | 6 [4—10] | 6 [4—8] | — |
| **Cardiac Disease** | | | | |
| Hypertension | Rate % (N) | 46.2% (212) | 34.0% (34) | 0.027 |
| Ischaemic Heart Disease | Rate % (N) | 23.5% (108) | 20.0% (20) | 0.51 |
| Atrial Fibrillation | Rate % (N) | 18.1% (83) | 11.0% (11) | 0.1 |
| Left Ventricular Failure | Rate % (N) | 11.5% (53) | 3.0% (3) | 0.009 |
| **Respiratory Disease** | | | | |
| COPD | Rate % (N) | 30.7% (141) | 24.0% (24) | 0.23 |
| Asthma | Rate % (N) | 9.6% (44) | 12.0% (12) | 0.46 |
| **Malignancy and Immunosuppressive Conditions** | | | | |
| Solid Organ Cancer | Rate % (N) | 15.5% (71) | 11.0% (11) | 0.28 |
| Haematological Malignancy | Rate % (N) | 9.6% (44) | 8.0% (8) | 0.71 |
| Autoimmune Disease | Rate % (N) | 7.6% (35) | 7.0% (7) | 1 |
| Chemotherapy | Rate % (N) | 3.9% (18) | 2.0% (2) | 0.55 |
| Corticosteroid use | Rate % (N) | 4.1% (19) | 3.0% (3) | 0.78 |
| On Immunosuppressants | Rate % (N) | 4.4% (20) | 1.0% (1) | 0.15 |
| **Vaccination status #** | | | | |
| PPV23 Vaccination | None % (N) | 51.4% (236) | 65.0% (65) | 0.0028 |
| | Under 6 months % (N) | 4.4% (20) | 8.0% (8) | |
| | Over 6 months % (N) | 44.2% (203) | 27.0% (27) | |
| Influenza Vaccination | None % (N) | 48.1% (221) | 54.0% (54) | 0.34 |
| | Under 6 months % (N) | 42.7% (196) | 35.0% (35) | |
| | Over 6 months % (N) | 9.2% (42) | 11.0% (11) | |
| **Admission Observations and Clinical Features** | | | | |
| Respiratory rate ($min^{-1}$) | Median [IQR] | 28 [24—32] | 30 [24—34] | 0.12 |
| Oxygen Sats (%) | Median [IQR] | 88 [84—91] | 86 [83—90] | — |
| Confusion | Rate % (N) | 28.8% (132) | 30.0% (30) | 0.81 |
| Hypotensive at admission | Rate % (N) | 14.8% (68) | 20.0% (20) | 0.22 |
| **Treatment** | | | | |
| Non-invasive ventilation | Rate % (N) | 7.6% (35) | 2.0% (2) | 0.044 |
| Endotracheal intubation | Rate % (N) | 17.6% (81) | 11.0% (11) | 0.14 |
| Tracheostomy | Rate % (N) | 4.4% (20) | 6.0% (6) | 0.44 |
| Inotropes required | Rate % (N) | 17.4% (80) | 9.0% (9) | 0.035 |

* Significance determined using Fisher's exact test (categorical variables) or not calculated due to missing values, 2 sample Wilcoxon Rank Sum test (continuous variables). Normality of distributions determined using Anderson-Darling normality test

‡ Index of deprivation was calculated using postcode to derive Lower-layer Super Output Area (LSOA) data from The Office of National Statistics (ONS)

### Vaccination status for pneumococcal and seasonal influenza was verified for each patient from GP records.

**Supplementary Table 4: Annual population and hospital admissions**

Population size within the Bristol, North Somerset and South Gloucestershire CCG and Bath & NE Somerset CCG as based on data obtained from the Office for National Statistics (ONS). Number of adult patients seen are listed as a combined total for all 3 NHS Trusts, from data provided by NHS Digital. Data for patients seen in Emergency Departments is derived from Hospital Accident and Emergency Activity. Data for admissions is collated from Monthly Hospital Episode Statistics for Admitted Patient Care, Outpatient and Accident and Emergency data

| **Characteristic – N** | 2006 | 2007 | 2008 | 2009 | 2010 | 2011 | 2012 | 2013 | 2014 | 2015 | 2016 | 2017 | 2018 |
|---|---|---|---|---|---|---|---|---|---|---|---|---|---|
| **Population** | | | | | | | | | | | | | |
| Whole population | 1,031,918 | 1,040,674 | 1,047,210 | 1,053,791 | 1,028,596 | 1,070,120 | 1,080,886 | 1,131,336 | 1,104,211 | 1,118,820 | 1,131,336 | 1,139,791 | 1,140,236 |
| ≥ 16 y | 823,298 | 833,548 | 839,310 | 844,186 | 866,522 | 887,470 | 925,556 | 893,556 | 902,950 | 915,438 | 925,566 | 933,304 | 940,293 |
| 16-34 y | 237,508 | 287,368 | 288,798 | 288,536 | 297,243 | 306,361 | 301,317 | 301,317 | 305,932 | 313,489 | 319,384 | 322,914 | 323,715 |
| 35-49 y | 222,704 | 224,970 | 225,290 | 225,944 | 227,259 | 220,217 | 219,883 | 219,883 | 217,895 | 217,136 | 216,015 | 215,618 | 215,987 |
| 50-64 y | 176,998 | 178,690 | 179,822 | 180,714 | 184,550 | 186,294 | 185,960 | 185,960 | 188,442 | 191,215 | 194,082 | 196,991 | 197,565 |
| 65-79 y | 106,488 | 107,636 | 109,918 | 112,516 | 118,948 | 129,021 | 140,629 | 133,018 | 136,428 | 138,803 | 140,629 | 141,792 | 142,895 |
| ≥80 y | 34,094 | 34,884 | 35,487 | 36,476 | 38,522 | 45,577 | 55,456 | 53,378 | 54,253 | 54,795 | 55,456 | 55,989 | 60,131 |
| **Adults Admissions** | | | | | | | | | | | | | |
| Accident & Emergency | 266,411 | 261,328 | 270,012 | 266,714 | 268,253 | 271,948 | 279,953 | 286,314 | 275,960 | 28,3137 | 263,566 | 269,868 | 271,328 |
| Unplanned admissions | 103,590 | 96,665 | 123,004 | 105,774 | 108,151 | 107,704 | 106,753 | 109,235 | 114,415 | 120,960 | 126,315 | 133,640 | 137,521 |
| RR admissions * | 1.00 | 0.93 | 1.19 | 1.02 | 1.04 | 1.04 | 1.03 | 1.05 | 1.10 | 1.17 | 1.22 | 1.29 | 1.33 |
| **Tests Conducted** | | | | | | | | | | | | | |
| Blood Cultures | 41,975 | 42,586 | 42,033 | 43,735 | 44,983 | 46,620 | 46,021 | 47,955 | 48,336 | 50,028 | 50,852 | 51,186 | 52,904 |
| Urine Antigen | 52 | 143 | 308 | 960 | 1,591 | 1,909 | 2,141 | 2,251 | 2,645 | 3,517 | 3,535 | 3,706 | 3,405 |
| RR testing * | 1.00 | 1.02 | 1.01 | 1.06 | 1.11 | 1.15 | 1.15 | 1.19 | 1.21 | 1.27 | 1.29 | 1.31 | 1.34 |

* The relative ratio of unplanned hospital admissions and microbiological testing (blood culture and urine antigen combined) were calculated using 2006 as the reference year

**Supplementary Data 5: Box plot of age of adults hospitalised with parapneumonic effusions and all medical admissions 2006-2018**

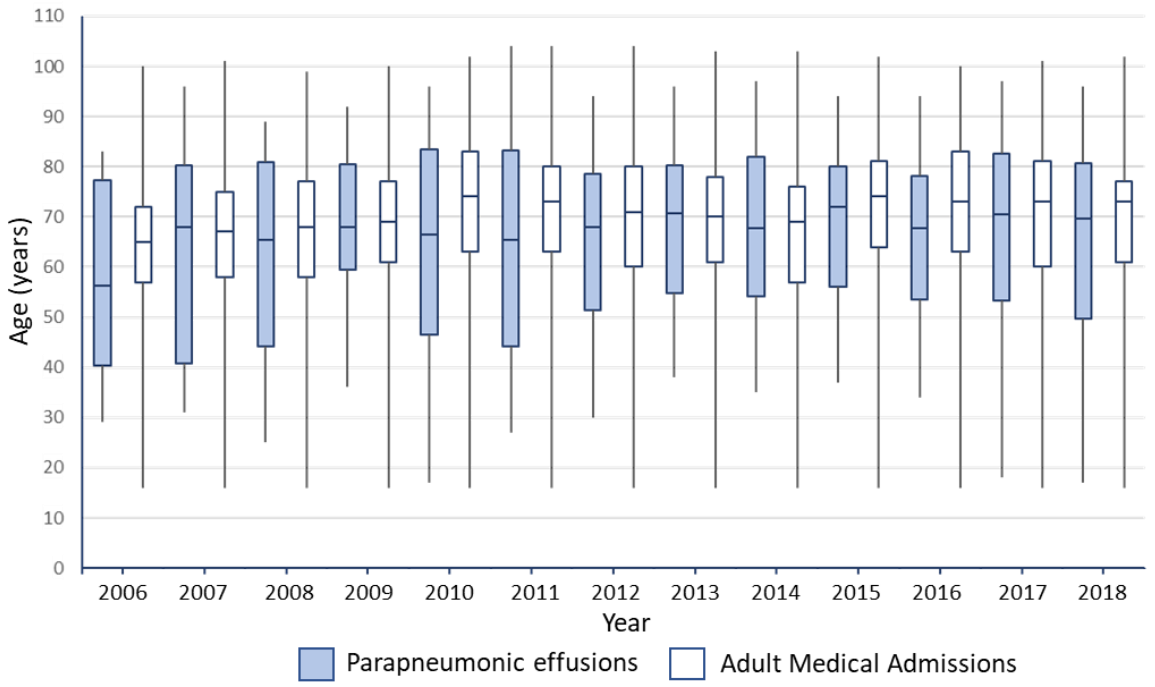

**Supplementary Data 6: Pleural disease attributable to significant serotypes throughout the study**

The percentage of pleural disease caused by significant serotypes from (A) PCV7, (B) PCV13-7 and (C) non-vaccine serotype groups, as a percentage of the total known serotype disease caused within year group

(A) PCV7 serotypes

| Year | 4 | 6B | 14 | 19F | 23F |
|---|---|---|---|---|---|
| 2006-2008 | 2.5 | 7.5 | 25.0 | 2.5 | 5.0 |
| 2009-2011 | 2.7 | 2.7 | 2.7 | 1.3 | 4.0 |
| 2012-2014 | 0.0 | 0.0 | 2.2 | 2.2 | 0.0 |
| 2015-2018 | 0.0 | 0.0 | 1.6 | 0.0 | 0.0 |

(B) PCV13-7 serotypes

| Year | 1 | 3 | 6A | 7F | 19A |
|---|---|---|---|---|---|
| 2006-2008 | 12.5 | 10.0 | 10.0 | 0.0 | 5.0 |
| 2009-2011 | 13.3 | 13.3 | 1.3 | 12.0 | 10.7 |
| 2012-2014 | 6.5 | 13.0 | 0.0 | 0.0 | 0.0 |
| 2015-2018 | 6.6 | 16.4 | 0.0 | 0.0 | 0.0 |

(C) Non-vaccine serotypes

| Year | 8 | 12F | 22F |
|---|---|---|---|
| 2006-2008 | 0.0 | 2.5 | 2.5 |
| 2009-2011 | 4.0 | 4.0 | 6.7 |
| 2012-2014 | 8.7 | 8.7 | 10.9 |
| 2015-2018 | 16.4 | 14.8 | 9.8 |

**Supplementary Data 7: Characteristics of Patients with pleural infection, by survival following presentation**

| Characteristic - N (%) | All Parapneumonic effusions | | SPE | | Pleural Infection | |
|---|---|---|---|---|---|---|
| | Survived >90 days | Died within 90 days | Survived >90 days | Died within 90 days | Survived >90 days | Died within 90 days |
| | N=453 | N = 106 | N=365 | N = 94 | N = 86 | N = 14 |
| Gender | | | | | | |
| - Male | 228 (50) | 57 (56) | 181 (49) | 50 (53) | 45 (52) | 7 (50) |
| - Female | 225 (50) | 49 (46) | 184 (51) | 44 (47) | 41 (47) | 7 (50) |
| Age – mean (IQR) | 69 (52-89) | 78 (68-84) | 68 (50-80) | 80 (68-89) | 56 (44-71) | 73 (67-88) |
| Smoking Status | | | | | | |
| - Current smoker | 174 (38) | 28 (26) | 128 (35) | 26 (28) | 44 (51) | 3 (21) |
| - Ex smoker | 180 (40) | 61 (58) | 159 (43) | 52 (55) | 21 (24) | 9 (64) |
| Vaccination Status | | | | | | |
| - PneumoVax (PPV23) | 204 (45) | 56 (58) | 176 (48) | 47 (50) | 27 (32) | 9 (64) |
| - Seasonal Flu | 239 (53) | 39 (40) | 195 (53) | 37 (39) | 43 (50) | 5 (35) |
| Pneumococcal vaccine serotype | | | | | | |
| Unknown serotype | 282/453 (62) | 55/106 (52) | 231/365 (63) | 50/94 (53) | 50/86 (58) | 5/14 (35) |
| PCV13 vaccine serotype | 95/181 (52) | 31/57 (54) | 65/143 (45) | 26/50 (52) | 30/36 (83) | 5/9 (56) |
| Non-vaccine serotype (NVT) | 16/181 (8) | 6/57 (11) | 14/143 (10) | 5/50 (10) | 2/36 (5) | 1/9 (11) |
| CPE/Empyema features | | | | | | |
| - Pus | 59 (13) | 0 (0) | - | - | 59 (66) | 0 (0) |
| - Loculation | 39 (9) | 10 (10) | - | - | 39 (45) | 10 (71) |
| - Fibrinolytic treatment | 27 (6) | 6 (6) | 0 (0) | 0 (0) | 27 (31) | 6 (43) |
| - Surgical Management | 29 (6) | 0 (0) | 0 (0) | 0 (0) | 29 (34) | 0 (0) |
| Co-morbidities | | | | | | |
| Chronic Resp Disease | 166 (37) | 54 (51) | 130 (36) | 49 (52) | 36 (42) | 5 (36) |
| COPD | 123 (27) | 44 (42) | 104 (28) | 40 (42) | 19 (22) | 4 (29) |
| Asthma | 48 (11) | 9 (8) | 35 (9) | 9 (10) | 13 (15) | 0 (0) |
| Other Chronic Resp Disease | 57 (10) | 11 (10) | 50 (13) | 9 (10) | 7 (8) | 2 (14) |
| Chronic Cardiac Disease | 226 (50) | 74 (69) | 190 (52) | 68 (72) | 36 (42) | 6 (43) |
| Hypertension | 187 (41) | 57 (54) | 156 (43) | 59 (63) | 31 (36) | 4 (33) |
| Ischaemic Heart Disease | 89 (19) | 38 (36) | 71 (19) | 39 (41) | 18 (21) | 1 (7) |
| AF | 63 (14) | 27 (25) | 59 (16) | 23 (24) | 4 (5) | 4 (29) |
| LVF | 42 (9) | 13 (12) | 39 (11) | 13 (14) | 3 (3) | 0 (0) |
| Other CVS | 16 (4) | 2 (2) | 16 (4) | 5 (5) | 0 (0) | 2 (14) |
| Immunosuppression | | | | | | |
| Autoimmune Disease | 24 (5) | 8 (8) | 16 (4) | 7 (7) | 8 (9) | 1 (7) |
| Oral Corticosteroids | 18 (4) | 4 (4) | 16 (4) | 3 (3) | 2 (2) | 1 (7) |
| Other immunosuppressive med | 17 (4) | 6 (6) | 14 (4) | 5 (5) | 3 (3) | 1 (7) |
| Solid organ cancer | 58 (13) | 24 (23) | 49 (13) | 21 (21) | 9 (10) | 3 (21) |
| Haematological malignancy | 38 (8) | 13 (12) | 34 (9) | 10 (11) | 4 (5) | 3 (21) |
| Recent chemotherapy | 15 (3) | 5 (5) | 14 (4) | 4 (4) | 1 (1) | 1 (7) |
| Other Risk Factors | | | | | | |
| Upper GI/Swallow problem | 168 (37) | 54 (52) | 136 (37) | 48 (51) | 29 (34) | 6 (43) |
| Chronic Liver Disease | 17 (4) | 8 (8) | 14 (4) | 7 (7) | 3 (3) | 1 (7) |
| Diabetes Mellitus | 81 (18) | 16 (15) | 66 (18) | 16 (17) | 15 (17) | 0 (0) |
| CVA disease | 24 (5) | 13 (12) | 19 (5) | 10 (11) | 5 (6) | 3 (21) |

| | | | | | | |
|---|---|---|---|---|---|---|
| Alcohol misuse | 41 (9) | 11 (10) | 29 (8) | 9 (10) | 12 (14) | 2 (14) |
| Intravenous drug usage | 16 (3) | 8 (8) | 10 (3) | 7 (7) | 6 (7) | 1 (7) |
| Obesity or BMI<17 | 16 (3) | 4 (4) | 9 (3) | 4 (4) | 7 (8) | 0 (0) |
| Admission Features | | | | | | |
| Length of Stay – median (IQR) | 11 (7-23) | 14 (6-24) | 10 (5-19) | 10 (4-21) | 17 (11-24) | 18 (11-28) |
| ICU/HDU care | 93 (20) | 20 (20) | 75 (20) | 18 (21) | 18 (21) | 2 (14) |
| - Intubation and ventilation | 73 (16) | 20 (20) | 62 (17) | 18 (21) | 11 (13) | 2 (14) |
| - Inotropic support | 71 (15) | 21 (21) | 62 (17) | 19 (22) | 9 (10) | 2 (14) |

**Supplementary Data 8: Testing of proportional hazards in Cox Regression model**
(A) Log-Log(survival) curve plot of SPE (red line) and pleural infection (blue line) cases in the cohort. Scaled Schoenfeld residuals are shown for (B) age and (C) loculation, demonstrating log hazard-ratio function is constant over time as gradient of each slope is zero.

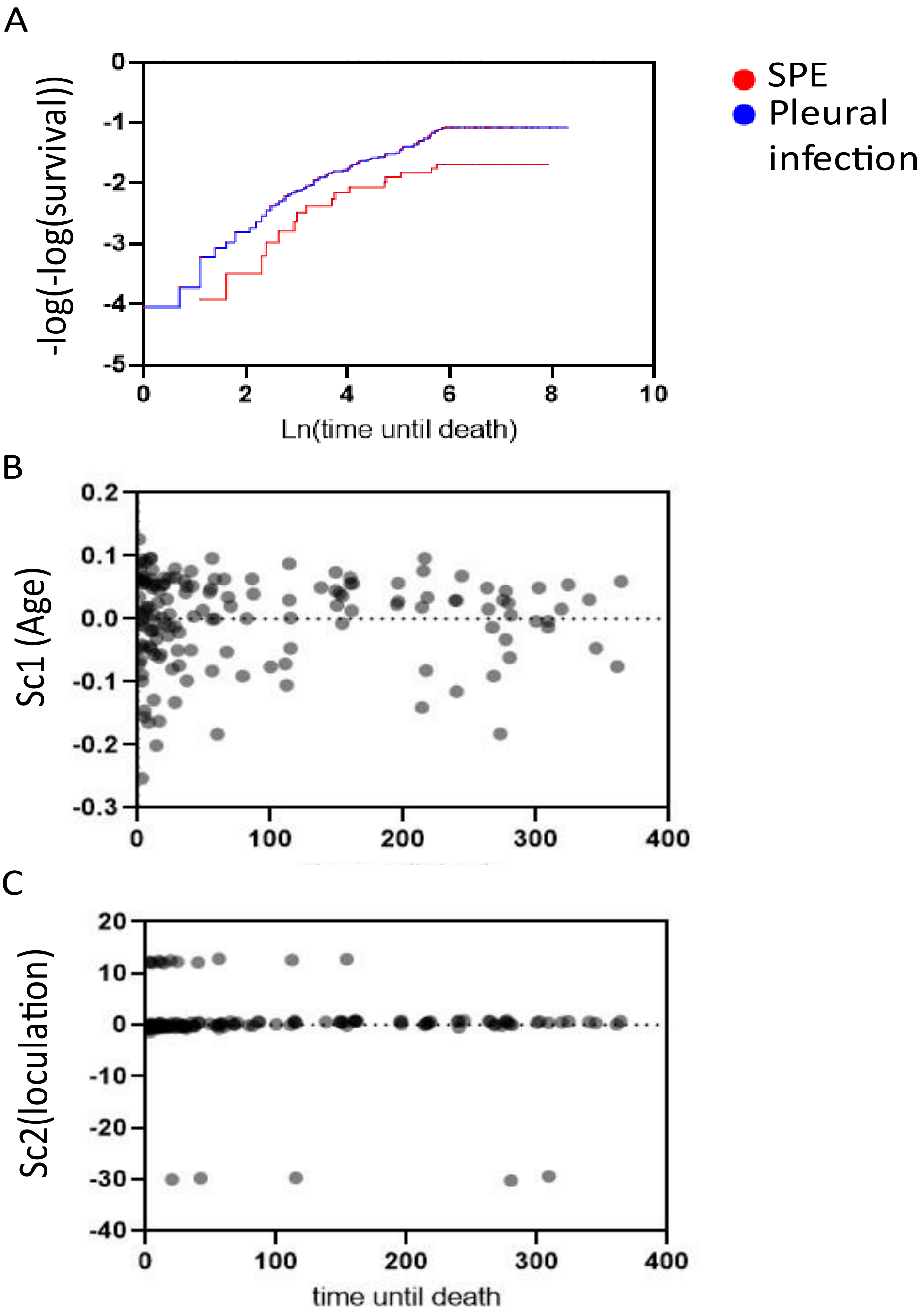

**Supplementary Data 9: Table of Abbreviations and Definitions**

| Abbreviation | Explanation |
|---|---|
| AF | Atrial Fibrillation |
| AMTS | Abbreviated Mental Test Score |
| BMI | Body Mass Index |
| CAP | Community Acquired Pneumonia |
| COPD | Chronic Obstructive Pulmonary Disease |
| CPE | Complex Parapneumonic Effusion |
| CVA | Cerebrovascular Accident |
| CVS | Cardiovascular System |
| ETT | EndoTracheal Tube |
| HIV | Human Immunodeficiency Virus |
| HDU | High dependency unit |
| IHD | Ischaemic Heart Disease |
| IPD | Invasive Pneumococcal Disease |
| IQR | Interquartile Range |
| ITU | Intensive Therapy Unit |
| LRTI | Lower Respiratory Tract Infection |
| LSOA | Lower-layer Super Output Area |
| LVF | Left Ventricular Failure |
| MALDI-TOF | Matrix-assisted laser desorption/ionisation/time of flight |
| ONS | The Office of National Statistics |
| PCV | Polysaccharide conjugate vaccine |
| PHE | Public Health England |
| PPV23 | Pneumococcal Polysaccharide Vaccine |
| RAPID | Renal, Age, Purulence, Infection source, & Dietary factors Score |
| SD | Standard Deviation |
| SPE | Simple Parapneumonic Effusion |
| ST | Serotype |
